# ACE-Neuro: A Tailored Exercise Oncology Program for Neuro-Oncology Patients – Study Protocol

**DOI:** 10.1101/2021.09.08.21263307

**Authors:** Julia T. Daun, Lauren C. Capozzi, Gloria Roldan Urgoiti, Meghan H. McDonough, Jacob C. Easaw, Margaret McNeely, George J. Francis, Tanya Williamson, Jessica Danyluk, Emma McLaughlin, Paula A. Ospina, Marie de Guzman Wilding, Lori Radke, Amy Driga, Christine Lesiuk, S. Nicole Culos-Reed

## Abstract

**Background:** Patients with primary brain tumours lack access to exercise oncology and wellness resources. The purpose of the Alberta Cancer Exercise – Neuro-Oncology (ACE-Neuro) study is to assess the feasibility of a tailored neuro-oncology exercise program for patients across Alberta, Canada. The primary outcome is to assess the feasibility of ACE-Neuro. The secondary outcome is to examine preliminary effectiveness of ACE-Neuro on patient-reported outcomes and functional fitness.

**Methods:** Neuro-oncology patients with a malignant or benign primary brain tumour that are pre, on, or completed treatment, are >18 years, and able to consent in English are eligible to participate in the study. Following referral from the clinical team to cancer rehabilitation and the study team, participants are triaged to determine their appropriateness for ACE-Neuro or other cancer rehabilitation or physiatry resources. In ACE-Neuro, participants complete a tailored 12-week exercise program with pre-post assessments of patient-reported outcomes, functional fitness, and physical activity. ACE-Neuro includes individual and group-based exercise sessions, as well as health coaching.

**Conclusion:** We are supporting ACE-Neuro implementation into clinical cancer care, with assessment of needs enabling a tailored exercise prescription.

## Introduction

While there is evidence supporting the role of exercise and physical activity (PA) for all individuals living with cancer [1], certain tumour groups, including patients with primary brain tumours (i.e., neuro-oncology), are underrepresented in this literature [2]. Primary brain tumours are defined as tumours that start in the brain cells and rarely spread outside of the central nervous system [3]. Patients with primary brain tumours are often presented with poor survival prognoses and undergo intensive treatments that result in cognitive and physical impairments, impacting activities of daily living (e.g., speech, balance, coordination), as well as quality of life [2, 4-6]. In Canada, Glioblastoma Multiforme (GBM) is the most commonly diagnosed brain cancer in adults [7]. As an advanced cancer population with a median survival of 12-14 months, and 5-year survival rate of 1% for adults over 55,[8] supporting patients to engage in exercise and physical activity may aid in supporting wellness and enhancing quality of life. To optimize potential effectiveness, a multidisciplinary collaboration across the medical, rehabilitation, and exercise specialist teams to enhance access to tailored PA resources is essential [2, 9, 10].

Exercise work to date in neuro-oncology has been limited, with the few studies supporting exercise feasibility and potential impacts, including decreasing symptom burden and improving physical function, cardiorespiratory fitness, cognition, quality of life, and emotional well-being [11] [12] [13]. Given the early state of this literature, work must continue to assess the role of exercise for individuals with brain tumours, and in particular assess the feasibility of implementation into clinical care and how to best tailor exercise based on the unique needs and significant treatment-related side effects that remain a major burden and negatively impact quality of life in this patient population [14-17].

Within Alberta, we have implemented the Alberta Cancer Exercise (ACE) program [18] over the past five years, and effectiveness is currently being assessed in a dataset of over 2300 participants. However, ACE primarily includes participants from breast, prostate, and colorectal tumour groups. Thus, there remains a critical need for clinical workflows to support building exercise referral into the cancer care system specifically for underserved populations, such as neuro-oncology. Building from ACE, and with a focus on co-creation of tailored programming with patients, clinicians, and researchers, our work aims to: (1) provide a tailored exercise program for neuro-oncology patients, considering addressing needs earlier in the care pathway, from diagnosis through treatment and into longer term survivorship; (2) provide models of delivery of exercise oncology programs to enhance access (i.e., remote delivery, home support, individual vs group); and (3) to build this improved access systematically within the neuro-oncology clinics in Calgary and Edmonton, to ensure that all patients diagnosed with brain tumours can access wellness resources during their cancer care journey.

The primary outcome of this work is to assess the feasibility of a tailored neuro-oncology exercise program for patients (i.e., ACE-Neuro-Oncology; ACE-Neuro), being treated at the two tertiary cancer centres in Alberta – the Tom Baker Cancer Centre (TBCC) in Calgary, and the Cross Cancer Institute (CCI) in Edmonton. Feasibility includes rates of referral and enrolment, program adherence, measurement completion, and adverse event reporting. Specific outcomes related to the rehabilitation triage clinic will be reported separately. Secondary outcomes are to examine the preliminary effectiveness of the neuro-oncology exercise program on patient-reported outcomes, functional fitness, and physical activity levels. We hypothesize that ACE-Neuro will be feasible, with ≥50% eligible patients referred to ACE-Neuro, ≥50% of those enrolled will complete the intervention, ≥60% of those who complete the intervention will complete pre- and post-intervention measures, ≥40% of those who complete the intervention will complete follow-up measures, and no major adverse events will occur. We also hypothesize that ACE-Neuro will be effective, as measured by improvements in patients’ physical and psychosocial well-being as well as physical activity levels (individual level outcomes), and a more integrated workflow in the clinical cancer care setting that includes exercise as part of standard clinical practice (systems level outcome).

## Methods

### Study Design and Procedure

This study was approved by the University of Calgary Health Research Ethics Board of Alberta (HREBA) – Cancer Committee (CC) - HREBA.CC-20-0322. Using the successful implementation model of the exercise oncology program developed in ACE, the proposed feasibility study includes a neuro-oncology cohort within a mixed methods study design.

### Participants

All neuro-oncology patients with a malignant or benign primary brain tumour that are pre, on, or completed treatment in Alberta, Canada, are >18 years, and able to consent in English are eligible to participate in the study. Recruitment began in April 2021 and is expected to close in Spring 2023, with follow-up assessments concluding a year later (Spring 2024). Because the main outcome of this study is feasibility, no *a priori* sample size has been calculated. Based on current clinical numbers, and previous work done with neuro-oncology patients at CCI, we anticipate approximately 25-30 eligible patients per year, per site.

### Recruitment & Referral

The study flow is depicted in Figure 1. Our aim is to support referral of eligible neuro-oncology patients to ACE-Neuro. Recruitment procedures are dependent on the site. Within Calgary (i.e., TBCC), the clinical team will send a referral to rehabilitation oncology via Alberta’s *Putting Patients First Questionnaire* in the electronic oncology information system. The clinical team, based on their judgment, may not refer patients they deem to be ineligible, for reasons such as disease status, not interested, unable to participate in exercise, do not speak English, or other clinical reasons. In Edmonton (i.e., CCI), neuro-oncology patients will be introduced to ACE-Neuro during their usual triage assessment that is conducted by an occupational therapist. Patients will be provided with a study brochure and instructed to contact the study team.

**Figure 1.**
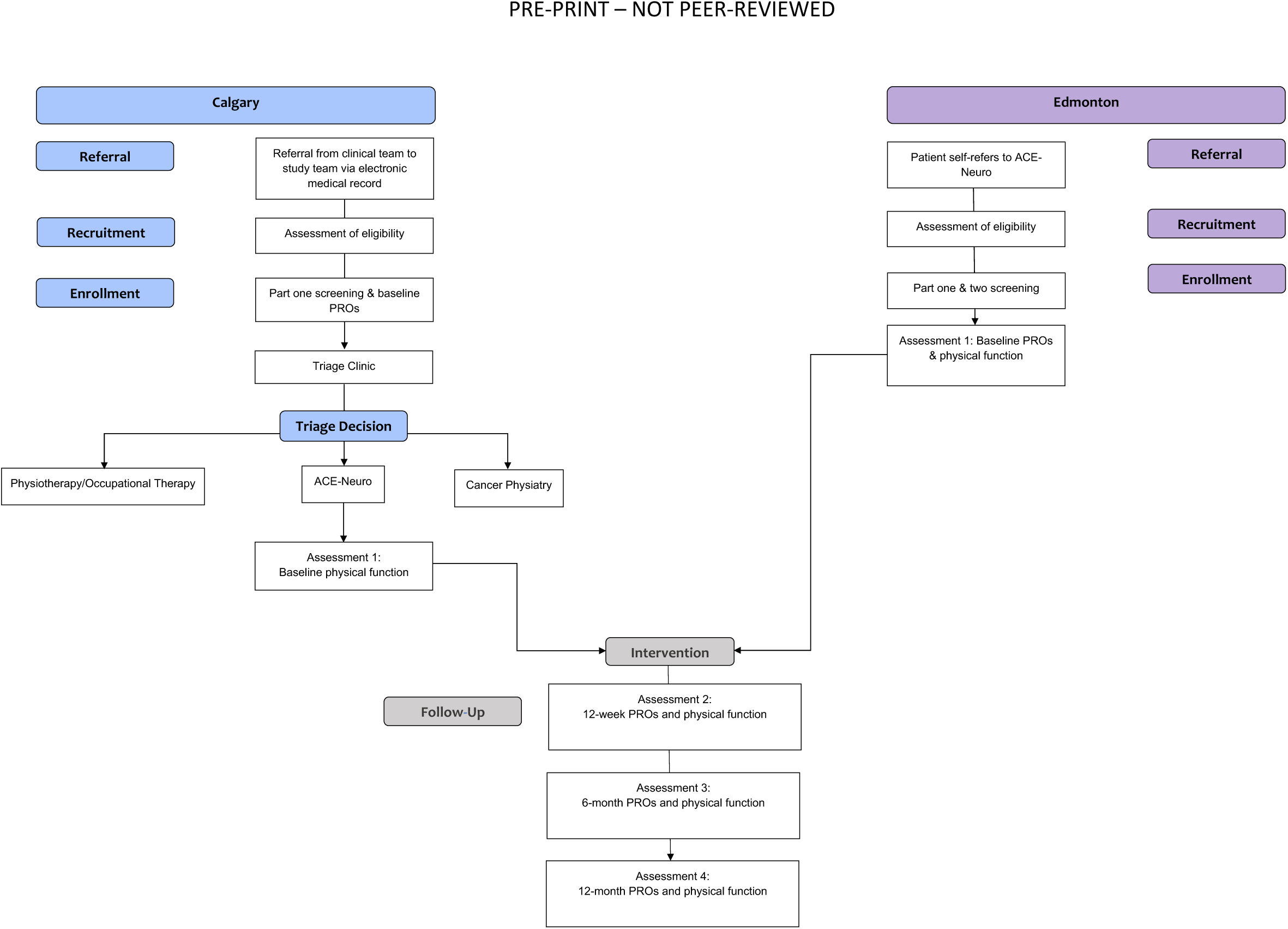
Flow Diagram for the Multi-Site Single-Arm ACE-Neuro Study. Recruitment began in April 2021. Follow-up assessments are expected to conclude in Spring 2024. PROs = patient-reported outcomes

**Figure 2.**
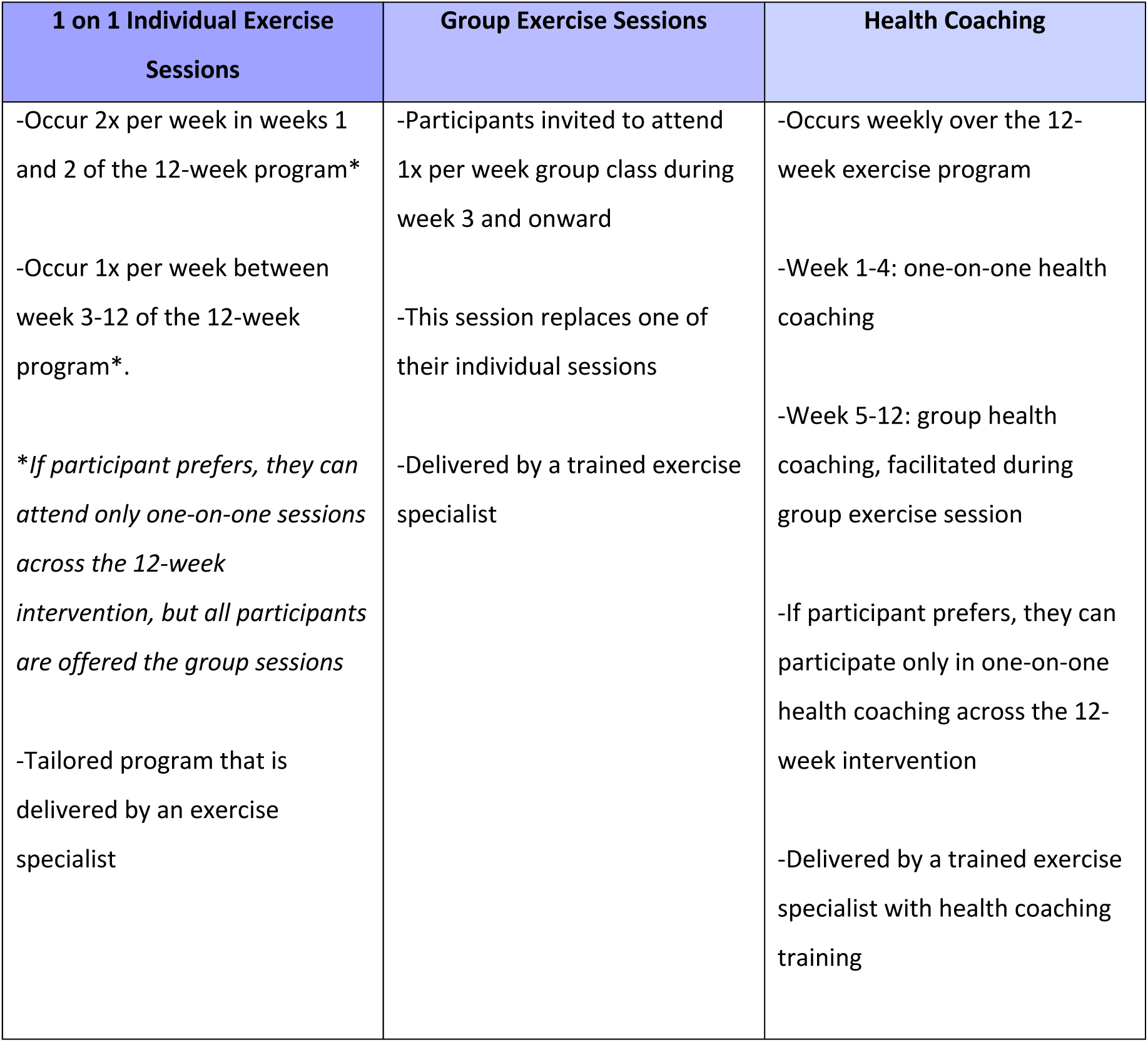
Overview of Exercise Intervention Components.

Once referred to ACE-Neuro, the study coordinator at each site contacts potentially eligible patients and presents a full introduction to the study. Patients that agree to participate are sent the study consent form via REDCap, a secure web application (Research Electronic Data Capture; REDCap) [19]. After consenting to the study, all patients undergo a two-part screening procedure prior to beginning the exercise program. In Calgary, this procedure includes the following:

1. **PART ONE SCREENING**: Patients will complete health and medical history screening, including a Health History Questionnaire, an Identifying Information Questionnaire (i.e., demographics), and the Physical Activity Readiness Questionnaire; PAR-Q+. In addition, patients complete baseline patient-reported outcomes (PROs), which are further outlined below. This screening and all questionnaires are completed via REDCap.
2. **PART TWO SCREENING**: Cancer Physiatry (i.e., physical medicine and rehabilitation) is established at the TBCC, thus patients will attend a Neuro Oncology Rehabilitation Triage Clinic, led by a Resident Physician and the ACE-Neuro Study Coordinator, to assess the patients’ readiness for participating in ACE-Neuro. During this 45-minute appointment, the medical and functional history is reviewed, a full neurological examination is performed, and he Short Physical Performance Batter screening test is performed [20]. From this, Karnofsky performance and Eastern Cooperative Oncology Group (ECOG) scores are determined. Patients will be triaged to ACE-Neuro, Cancer Physiatry, Rehabilitation Oncology (i.e., Physiotherapy/Occupational Therapy), or a combination of these services. If not initially triaged to ACE-Neuro, patients will be re-referred to the ACE-Neuro study team once deemed appropriate by their clinical team. During COVID-19, additional Alberta Health Services-regulated screening procedures will take place in advance of this in-person appointment.

In Edmonton, Cancer Physiatry is not part of the cancer care system. Thus, as part of usual care, patients will be assessed by an occupational therapist, which includes completing the Short Physical Performance Battery screening test. Following self-referral to ACE-Neuro and participant consent to the study, the subsequent screening procedure includes:

1. **PART ONE SCREENING**: A Clinical Exercise Physiologist will complete health and medical screening for participants using the Health History Questionnaire, Identifying Information Questionnaire, and the Physical Activity Readiness Questionnaire; PAR-Q+.
2. **PART TWO SCREENING**: The Clinical Exercise Physiologist will obtain physician approval for participation in ACE-Neuro.

### The Exercise Intervention

#### Exercise Sessions

Depending on COVID-19 restrictions and participant preferences, the intervention will be delivered remotely (i.e., via Zoom), or in-person (i.e., at the University of Calgary or University of Alberta cancer and exercise-specific facilities). Upon entering the study, participants will be provided with a welcome package including an overview of the 12-week program, the role of exercise for neuro-oncology, understanding the FITT (Frequency, Intensity, Time, and Type) principle, instructions for using the activity tracker, educational topics and their respective handouts, and additional resources, including a <u>Cancer and Exercise Wellness Manual</u>. The 12-week exercise intervention will be tailored to each participant, with programs designed by an exercise specialist. Following published guidelines [1], and the established ACE [18], program protocol, the program will include twice-weekly supervised exercise sessions led by the study exercise specialist. Session intensity will be based on participants’ acute perceptions of energy and fatigue and monitored using Borg’s Rating of Perceived Exertion (RPE; 1-10) scale [21]. Sessions will be 30-60 minutes, tailored to meet the unique needs of each individual and progressed over time, and overall may include the following: 5-10-minute warm-up focusing on mobility and light aerobic movements (RPE ∼1-3); a 15-40-minute aerobic, resistance, and balance training circuit (RPE ∼2-6); followed by a 10-15-minute cool down, including flexibility training (RPE ∼1-2). After 2 weeks of the individual sessions, participants will be offered a once-weekly group in-person or virtual (depending on COVID restrictions) session with other ACE-Neuro participants. This group session will replace one of their weekly individual sessions, and is designed to foster social connections that are central to the ACE model [18]. Finally, the exercise program will follow an “*exercise and educate*” framework that is based on motivational interviewing, health coaching, and health behaviour change [22, 23] that includes the education topics of (1) goal setting, (2) behaviour change, (3) stress management, (4) self-compassion, (5) sleep, and (6) social support. Education topics will be discussed every two weeks at the end of the exercise session, during cool-down. In addition, participants will have the option to attend a live webinar on each topic during their 12-week program.

#### Health Coaching

All participants will have the choice to participate in health coaching calls [22], provided by a health coach with exercise oncology specific training. Health coaching calls will take place weekly for 15-30 minutes following an individualized exercise training session, and will be delivered remotely (e.g., via end-to-end encrypted Zoom or phone call) at the participants’ preferred date and time. Health coaching calls will use a participant-centered approach, with consideration given to participant-determined discussion, self-discovery, and the coach-participant relationship [22]. Health coaching will be delivered one-on-one for the first month of the program (week 1-4), followed by group health coaching for the remainder of the 12 weeks (week 5-12), for the participants that attend the group-based classes.

To ensure consistency in both the health coaching and exercise program delivery across participants, and to ensure that the principles of health coaching are being followed during calls, fidelity checks will take place throughout the study. Ten percent of all calls and exercise sessions will be randomly selected for evaluation, and a standardized fidelity ‘checklist’ form will be completed by blind trained assessors (i.e., experts in the field; trained graduate students).

### Timeline of Assessments

In addition to attending the triage clinic, consenting patients will complete assessments at five timepoints: (1) baseline PROs and health screening, pre-triage clinic, (2) baseline physical function, post triage clinic, (3) post-program (twelve weeks), (5) six months, (6) twelve months (Figure 1). Each timepoint will include the completion of PROs via REDCap and the assessment of physical function (online via Zoom or in-person). Objective PA will only be collected during the intervention (baseline to twelve weeks).

### Study Measures

#### Demographics and Clinical Characteristics

Demographics and clinical characteristics will be collected via a secure web application (Research Electronic Data Capture; REDCap) [19] and confirmed with chart review by the Study Coordinator (JTD) and second author (LCC) in ARIA. Data collected will include diagnosis and treatment details, sex, self-selected ethnicity and gender, employment status, annual family income, smoking status, and alcohol consumption.

#### Primary Outcome: Feasibility

To assess feasibility, we will track referral rate, enrolment rate, program adherence, measurement completion rate, and adverse events. All feasibility aspects of the triage clinic will be reported separately. All feasibility thresholds are based on feedback from the clinical team and other feasibility work in exercise oncology [24] [25].

##### Referral to ACE-Neuro

To examine the feasibility of referral, the number of patients referred from the clinical team to ACE-Neuro will be tracked. The pre-determined threshold is ≥50%.

##### Enrollment into the Study

To examine the feasibility of enrollment, the number of patients that enrol into the study after hearing the full study introduction will be tracked. The pre-determined threshold is ≥50%.

##### Program Adherence

The number of participants that complete the exercise intervention will be tracked. The pre-determined threshold is ≥50%.

##### Measurement Completion

Measurement completion rate, defined as the percentage of completed measures (PROs, physical function, objective physical activity levels) will be tracked. The pre-determined measurement completion is ≥60% for pre-and post-intervention assessments, and ≥40% at the two follow-ups (6 and 12 months).

##### Adverse Events

To assess the safety of the intervention, all adverse events will be tracked and reported using a standardized adverse event reporting form, that ranks events as level 1 (minor incident with no lost time beyond day of injury; temporary, immediate care), level 2 (medical aid with no lost time beyond day of injury; medical care beyond first aid), and level 3 (serious injury or death).

#### Secondary Outcome: Preliminary Effectiveness

To examine the preliminary effectiveness of the exercise intervention, PROs and assessments of physical function will be conducted. All measures were chosen based on their established validity, previous use in cancer patients, and relevance to the evaluation of the benefits of an exercise oncology program.

##### Patient-Reported Outcomes

Patient-reported outcomes (PROs) will include symptom burden, physical activity levels, quality of life, cognitive function, and fatigue. Symptom burden will be assessed using the revised Edmonton Symptom Assessment System (ESAS-r), which evaluates nine common symptoms experienced by cancer patients [26, 27]. Self-reported exercise levels will be assessed using the modified Godin Leisure-Time Exercise Questionnaire (GLTEQ) [28] which reports mild, moderate, vigorous intensity and aerobic, resistance, and flexibility activities that last more than 10 minutes. Quality of life will be assessed using the Functional Assessment of Cancer Therapy-Brain (FACT-Br), which includes subscales for physical well-being, social/family well-being, emotional well-being, functional well-being, and additional neuro-oncology specific concerns, such as reporting seizures, speech difficulties, memory, etc. [29]. Cognition will be assessed using Functional Assessment of Cancer Therapy-Cognition (FACT-Cog), which includes subscales for perceived cognitive impairments [30]. Fatigue will be assessed using the Functional Assessment of Chronic Illness Therapy Fatigue Scale (FACIT-F)[31]. Finally, physical activity preferences will be collected from participants at baseline to help tailor their individual exercise prescription.

##### Assessment of Functional Fitness

Assessments of physical function will follow the set protocols within the larger ACE study [18], and are designed to be able to be completed in-person or via remote delivery (online assessment). All fitness assessments will be completed by an exercise specialist and will include assessments of body composition, muscular strength, muscular endurance, balance, flexibility, and cardiorespiratory fitness. Given the online nature for the start of ACE-Neuro, only the online assessments are indicated here. See Supplementary File 1 for a table of the in-person assessments. For online assessments, resting heart rate will be measured by the study-provided activity tracker or via manual palpation. Resting blood pressure will be measured if the participant has an at-home blood pressure monitor. Participants’ height and weight measurements from the triage clinic will be used. Muscular endurance will be measures by the 30 second sit-to-stand test [32, 33]. Static balance will be measured by the single-leg-stance following the Canadian Society for Exercise Physiology (CSEP) protocol [34, 35]. Flexibility will be measured using the sit and reach test [36, 37] and the shoulder flexion test [38]. Cardiorespiratory fitness will be measured using the 2-Minute Step Test [39].

##### Objective Physical Activity

Objective physical activity will be measured via the use of a consumer-level wrist-worn activity tracker (WAT; i.e., Garmin Vivofit 4). Garmin wearable activity trackers have high inter-device reliability of step count and are widely used across health research [40]. The Garmin activity tracker will be provided to all participants to objectively track PA habits throughout the intervention. Total weekly steps and PA minutes (i.e., mild, moderate, vigorous) will be tracked between baseline and week 12 of the program.

##### Qualitative Interviews and Photo Elicitation

Qualitative data will be gathered across the study timeline via interviews and photo elicitation [41], to inform the feasibility of ACE-Neuro, as well as to assess outcomes associated with participation in the ACE-Neuro program (i.e., benefits, barriers, satisfaction, impact on well-being, impact on sense of self). All participants will be invited to a 45-60-minute post-program interview, which will be recorded using: (1) a voice recorder if in-person or (2) end-to-end encrypted Zoom if remote. Participants will be interviewed within 2 weeks of completing the 12-week exercise program to limit recall bias, allowing them to reflect on their experience in the program. The interview guide is informed by the COM-B behaviour change framework examining participant *capabilities, opportunities, motivations*, and *behaviour* [23]. The qualitative phase of this study will be guided by an Interpretive Description methodology and constructivist philosophy [42]. Interpretive description is well-established and has been used to guide numerous health-disciplined qualitative papers [43-48]. With consent, participants who engage in this process will have candid photographs taken, and/or will be encouraged to take photos of their journey via their mobile devise/personal camera, or a study-provided disposable camera. Standardized instructions for capturing photos will be provided to participants. Photos will be sent to the study team via the secure, end-to-end encrypted messaging app, Signal (https://signal.org/). When available, photos will be presented to the patient during the interviews to elicit memories and feelings. This can be a powerful tool to reinforce the nature of their exercise oncology program experience, and is a valid tool for aiding more in-depth understanding of the patient experience [41]. Photos gathered from and/or taken of participants will be transferred from the Signal App, study-provided disposable camera, or from the study team and stored on a secure University of Calgary server.

#### Statistical Analysis

##### Quantitative Data

Descriptive characteristics of participants will be presented as mean ± standard deviation or percentages. Feasibility will be reported descriptively in relation to the pre-determined thresholds. We will investigate preliminary effectiveness of our secondary outcomes. Descriptive statistics will also be reported for feasibility numbers, PROs, functional fitness, and objective PA. Change scores will be calculated for PROs and physical function and to calculate power for a future fully-powered trial. Where available, the minimum clinically important difference (MCID) [49] will be reported as an indicator of clinical significance, which is appropriate for a pilot study.

##### Qualitative Data

Interviews will be transcribed verbatim via ExpressScribe and coded in NVivo 12. As per an Interpretive Description methodology, the data will be inductively analyzed by two independent authors who will generate themes from the codes, followed by critical feedback from experts in qualitative research and exercise oncology.

## Discussion

The purpose of this pilot study is to assess the feasibility of a tailored exercise oncology program, ACE-Neuro, for individuals with primary brain tumours. The primary outcome of this study is to determine the feasibility of referral, enrolment, program adherence, measurement completion, and adverse events. The secondary outcomes include examining preliminary effectiveness of the neuro-oncology exercise program on patient-reported outcomes, functional fitness, and physical activity levels.

While exercise oncology programming is available for all tumour groups, patients with primary brain tumours remain underrepresented in the research process and underserved in exercise resources. By delivering one-on-one sessions, we hope this work will provide additional opportunities to participants that may have struggled in group-based settings that did not fully address their needs.

Findings from this work will be disseminated through academic channels (e.g., manuscripts, presentations), as well as through non-academic leveraging (e.g., knowledge translation initiatives such as patient group presentations, online resources for patients). Using an implementation framework approach (i.e., the RE-AIM framework; Reach, Effectiveness, Adoption, Implementation, Maintenance) [50], findings will target existing programs to improve resources, program delivery, and fine-tune models of care [51].

## Limitations

First, due to the smaller population of neuro-oncology patients, we may not have the power to determine effectiveness of the intervention on secondary outcomes (i.e., PROs, physical function). Instead, the feasibility and preliminary effectiveness findings of this study will be used to calculate the power for a future exercise trial in this population.

Second, due to COVID-19, the delivery of our programming will initially be offered online, with the option of returning to in-person delivery once restrictions lift. Depending on participant preferences, we will continue to offer both options throughout the study duration. With the transition from online to in-person, we risk inconsistency in the results of our assessment of physical function, nevertheless, the assessments that have been chosen can be replicated in both settings with the same protocol in place.

## Conclusion

Patients with primary brain tumours are a clinically underserved patient population that are underrepresented in the exercise oncology research. To address this gap, ACE-Neuro is a tailored exercise oncology program that will be implemented into the clinical care pathway across Alberta. ACE-Neuro provides an opportunity to provide patient-centered supportive cancer care that enhances wellness for individuals living with a brain tumour diagnosis.

## Data Availability

De-identified data from this study are not available in a public archive. De-identified data from this study will be made available (as allowable according to institutional IRB standards) by emailing the corresponding author.

