## Supplementary Table 1 for "ACE-Neuro: A Tailored Exercise Oncology Program for Neuro-Oncology Patients – Study Protocol"

Supplementary File 1

Functional Fitness Assessment – In Person Assessment Overview

| | |
|---|---|
| Resting Vital Measures | Heart Rate<br>Blood Pressure |
| Anthropometric Measures | Height<br>*Measured using a Seca 217 Stadiometer*<br>Weight<br>*Measured using a 16 Health Carter Beam Scale* |
| Muscular Strength | Grip Strength<br>Measured using a hand dynamometer, following the Canadian Physical Activity, Fitness, and Lifestyle Approach (CPAFLA) |
| Muscular Endurance | 30-Second Sit-to-Stand |
| Balance | Single Leg Stance |
| Flexibility | Shoulder Flexion<br>*Measured using a goniometer*<br>Sit-and-Reach |
| Cardiorespiratory Fitness | 6-Minute Walk Test |
